# COVINet: A deep learning-based and interpretable prediction model for the county-wise trajectories of COVID-19 in the United States

**DOI:** 10.1101/2020.05.26.20113787

**Authors:** Yukang Jiang, Ting Tian, Wenting Zhou, Yuting Zhang, Zhongfei Li, Xueqin Wang, Heping Zhang

## Abstract

The cases of COVID-19 have been reported in the United States since January 2020. There were over 103 million confirmed cases and over one million deaths as of March 23, 2023. We propose a COVINet by combining the architecture of both Long Short-Term Memory and Gated Recurrent Unit and incorporating actionable covariates to offer high-accuracy prediction and explainable response. First, we train COVINet models for confirmed cases and total deaths with five input features, compare their Mean Absolute Errors (MAEs) and Mean Relative Errors (MREs) and benchmark COVINet against ten competing models from the United States CDC in the last four weeks before April 26, 2021. The results show that COVINet outperforms all competing models for MAEs and MREs when predicting total deaths. Then, we focus on the prediction for the most severe county in each of the top 10 hot-spot states using COVINet. The MREs are small for all predictions made in the last 7 or 30 days before March 23, 2023. Beyond predictive accuracy, COVINet offers high interpretability, enhancing the understanding of pandemic dynamics. This dual capability positions COVINet as a powerful tool for informing effective strategies in pandemic prevention and governmental decision-making.

## 1 Introduction

According to the New York Times [33], the early confirmed cases of COVID-19 were reported on January 21, 2020, in the United States. In March [40], the outbreak of COVID-19 was proclaimed as a “pandemic” by the World Health Organization. Since then, the United States has had the largest number of confirmed cases and deaths globally [24], where the confirmed cases and deaths were 103,910,087 and 1,135,344, respectively, as of March 23, 2023.

A vast majority of states in the United States issued a “stay at home” order to reduce the transmission of COVID-19 since March 2020 [18]. As the states are reopening to achieve normalcy, it is essential to predict the trajectories of COVID-19 based on actionable factors to provide the decision-makers with a quantitative and dynamic assessment. Here, we define the actionable factors as those that may be routinely surveilled and collected by the local and national authorities, such as the level of air pollution [34]. Among them, environmental factors affect the spread of infectious diseases. For instance, the hospitalization rate of H1N1 2009 had a disproportionate impact on high-poverty areas in New York City [4] and on the small population of racial/ethnic groups in Wisconsin [36]. Consequently, we consider county health ranking and roadmaps programs [32]. The details about the database are available from https://www.countyhealthrankings.org/reports/county-health-rankings-reports. We focus on health factors related to physical and social environments as well as demographics, which are selected based on variable importance ranking of the random forest, as summarized in Table 1.

**Table 1:** The list of five health factors related to their categories, meanings, and sources. The ranks of factors are the variable importance rankings of the random forest models for cumulative confirmed cases and deaths, respectively.

| Category | Factors<br>(Ranks) | Meanings | Sources |
| --- | --- | --- | --- |
| **Physical Environment** | Traffic volume (3, 3) | Average traffic volume per meter of major roadways in the county | Environmental Justice Screening and Mapping Tool |
|  | Severe housing problems (4, 4) | Percentage of households with at least 1 of 4 housing problems: overcrowding, high housing costs, lack of kitchen facilities, or lack of plumbing facilities | Comprehensive Housing Affordability Strategy (CHAS) data |
|  | Air pollution (6, 5) | Average daily density of fine particulate matter in micrograms per cubic meter (PM<sub>2.5</sub>) | Environmental Public Health Tracking Network |
| **Social & Economic Factors** | Some college (2, 1) | Percentage of adults ages 25-44 with some post-secondary education | American Community Survey, 5-year estimates |
| **Demographics** | Population (1, 2) | Resident population | Census Population Estimates |

There are many studies dedicated to forecasting the spread of COVID-19. The epidemic models are prevalent tools to predict the infection trajectories [23, 38, 41]. For example, the United States (US) COVID-19 Forecast Hub[14] is a data repository that collects and aggregates the predictions of various epidemic models for the US COVID data. Instead of relying on disease resumption, some authors proposed neural networks to precisely estimate the epidemic [20, 43]. These data-driven approaches had superior performance in predicting the dynamics of COVID-19. Yang et al. [43] proposed a Long Short-Term Memory (LSTM) [19] based model, and Bandyopadhyay and Dutta [5] compared three models, including LSTM, Gated Recurrent Unit (GRU) [11], and LSTM combined with GRU in predicting COVID-19. The LSTM combined with GRU had been proven to generate a high accuracy rate [8]. However, a deep learning-based model is generally complex and not useful in making informed decisions. Therefore, our primary goal is to build deep learning models that can help decision-making for the epidemic.

We propose COVINet, a model that utilizes LSTM and GRU networks to forecast disease dynamics at the county level. By incorporating three actionable features reflecting community health risk, as well as longitude and latitude data for each county, COVINet captures local impacts of the disease, identifies high-risk and low-risk factors, and provides valuable and actionable information for public health. To evaluate the performance of COVINet, we align our county-level results with the state-level predictions of ten competing models from the US Centers for Disease Control and Prevention (CDC) that used state-level data. Specifically, we aggregate our county-level results to match their scale for comparison. Additionally, after the prediction of the COVID-19 pandemic for all counties, we showcase our predictive model for the most severely affected county in each of the top 10 states with the highest number of confirmed cases, considering their paramount public health significance. Thus, COVINet’s interpretability sheds light on the “black box” of deep learning, providing a clear understanding of how actionable features impact the trajectory of the COVID-19 pandemic. Our work is to obtain accurate predictions in the projected trajectories of COVID-19 in the hot-spot areas and directly provide measurable and actionable responses to reduce the spread of COVID-19.

## 2. Methods

### 2.1. Data Sources

We collect the daily numbers of cumulative confirmed cases and deaths from January 21, 2020, to March 23, 2023, for infected counties in the US from the New York Times [33]. The daily cumulative confirmed cases and deaths are collected from health departments and the US CDC, where patients are identified as “confirmed” based on positive laboratory tests and clinical symptoms and exposure [33]. All risk factors are compiled from 2020 annual data on the County Health Rankings and Roadmaps program’s official website [32]. In addition, the longitude and latitude of each infected county are collected from Census TIGER 2000 [25]. Data analysis is conducted in Python 3.7 with TensorFlow-GPU 1.14.0 and Keras 2.3.0.

### 2.2. The selection of features

The input data are divided into two parts. The first part consists of the cumulative confirmed cases and deaths in the past fourteen days:

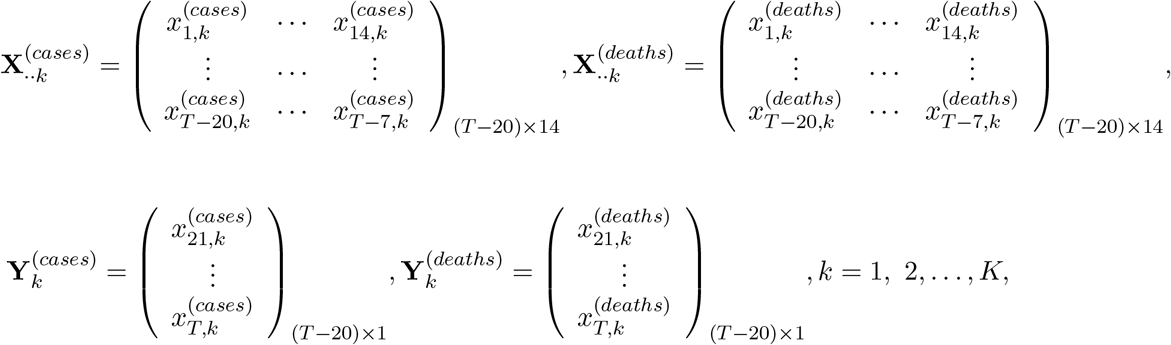

where *T* is the length of the training period, and *K* is the total number of coun-ties. 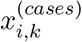 are the cumulative confirmed cases and 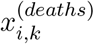 are the total deaths at the corresponding date. For example, *i* = 1 corresponds to the first day when the confirmed cases and deaths were officially reported. These cumulative confirmed cases and total deaths give rise to fourteen historical epidemic features as the first part of the input data. The other part of the inputs includes *J* county features, 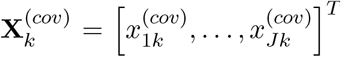. These features are three actionable factors in addition to the longitude and latitude of infected counties. Thus, *J* = 5 applies to the second part of our input data. Although the longitude and latitude of infected counties are not actionable features, we include them in our model because of their established importance in prediction [29, 31].

Our goal is to incorporate important features that can enhance the accuracy and interpretability of COVINet. To achieve this, we employ the random forest to screen the three actionable features. In a random forest, a common practice is to select features with the largest variances [8]. This approach selects the following three features: traffic volume, severe housing problems, and air pollution (PM_2.5_) (Table 2). Therefore, as presented in Figure 1, our proposed model uses nineteen features as the input data, comprising fourteen historical epidemic features and five county features (three selected actionable features, longitude, and latitude). Note that the input data are not predicted from the model.

**Table 2:** Comparison of the performance of COVINet and ten CDC models in predicting the disease dynamics using the MAE and MRE as the evaluation metrics. The results are reported for the top 10 states and all states in the US for a 7-day prediction. The results of COVINet have been averaged over 50 repetitions.

| Method | Top 10 States |  | All States |  |
| --- | --- | --- | --- | --- |
|  | MAE<sub>7</sub> | MRE<sub>7</sub> | MAE<sub>7</sub> | MRE<sub>7</sub> |
| COVINet(Proposed model) | **159.00** | **0.0049** | **49.48** | 0.0107 |
| UMass-MechBayes[17] | 163.05 | 0.0058 | 58.00 | 0.0079 |
| COVIDhub CDC-ensemble[30] | 167.88 | 0.0060 | 58.10 | **0.0077** |
| LANL-GrowthRate[26] | 173.55 | 0.0063 | 62.72 | 0.0080 |
| MOBS-GLEAM COVID[1] | 179.64 | 0.0065 | 66.37 | 0.0083 |
| COVIDhub-baseline [15] | 186.20 | 0.0067 | 71.64 | 0.0100 |
| IowaStateLW-STEM [37] | 187.25 | 0.0065 | 73.46 | 0.0082 |
| UT-Mobility[39] | 196.75 | 0.0072 | 72.33 | 0.0083 |
| CU-select[42] | 221.26 | 0.0074 | 82.07 | 0.0096 |
| CU-nochange[27] | 221.78 | 0.0075 | 82.18 | 0.0096 |
| JHU-IDD-CovidSP[22] | 409.73 | 0.0157 | 132.71 | 0.0216 |

**Figure 1:**
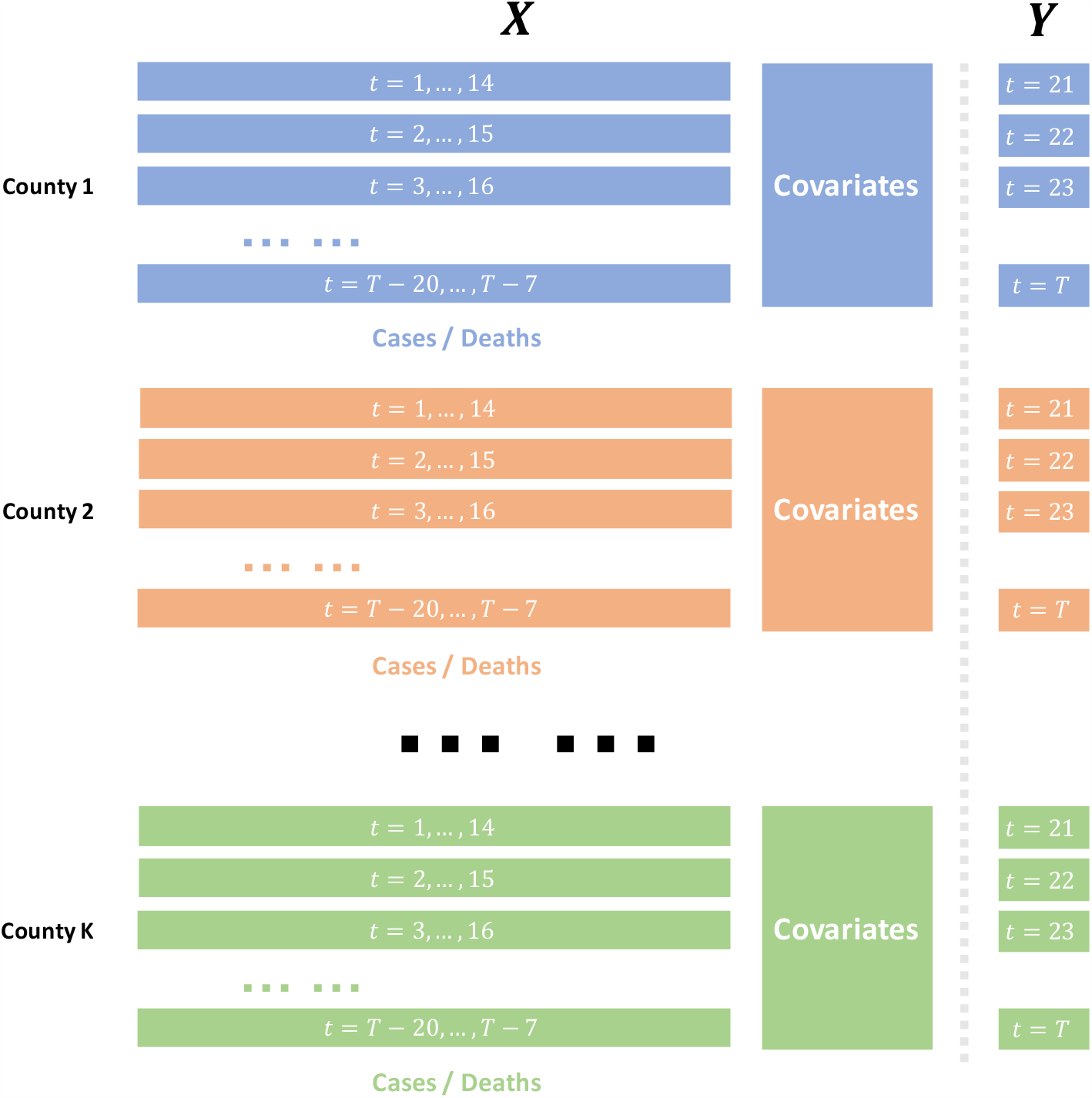
The structure of data usage in the models. Cumulative data for each county (confirmed or death cases) from the preceding 1st to 14th days serve as independent variables (*X*) for predicting the cumulative data (confirmed or death cases) as response variable *Y* on the 21st day. This process is repeated for subsequent days for each county. The method also integrates covariate data from different counties, collectively inputting them into the model, and conducts separate modeling for confirmed and death cases.

### 2.3. COVINet

#### 2.3.1. Model architecture

Our proposed model integrates an LSTM layer, a GRU layer [5, 11, 19], and a fully connected layer, formulated as:

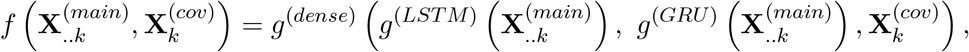

where *g*^(*dense*)^ is a fully connected layer, *g*^(*LST M*)^ is an LSTM layer, and *g*^(*GRU*)^ is a GRU layer. The time series of historical epidemic data 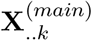 are the inputs of LSTM and GRU layers, which are typically used in time series analysis for the deep learning process. We then concatenate the outputs of these two layers, and the time-invariant county features 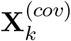 in a fully connected layer.

An LSTM layer (*g*^(*LSTM*)^) contains the input gate *in*_*t*_, the forget gate *f*_*t*_, the output gate *o*_*t*_, the cell state *c*_*t*_ (i.e., the hidden status), the candidate value 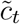, and the hidden state vector/final output *h*_*t*_. 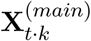 is a *t* ^*th*^ row of 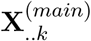 used as the input vector of the LSTM layer, then the iterative formula for each item is shown as follows:

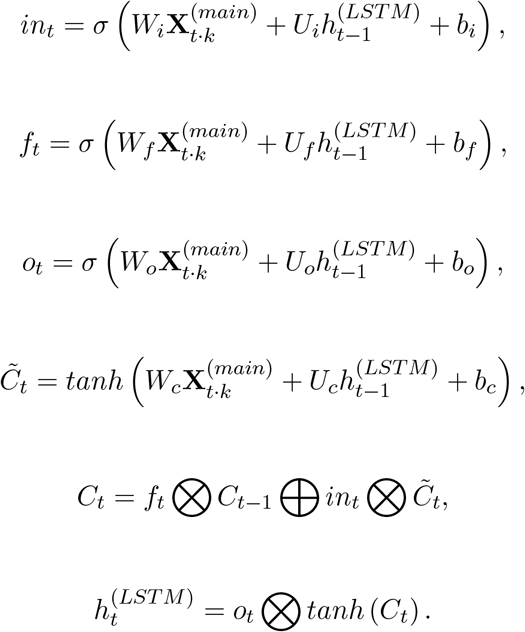

Comparatively, a GRU layer (*g*^(*GRU*)^) streamlines the operation. The layer removes the cell state *C*_*t*_, the information transmits in the hidden state (*h*_*t*_), input gate *in*_*t*_ and forget gate *f*_*t*_ emerge to form an updated gate *z*_*t*_, a reset gate *r*_*t*_ adds, and removes the final output gate. Thus, the corresponding update functions are:

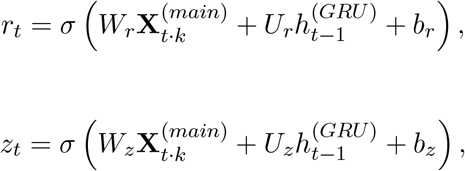

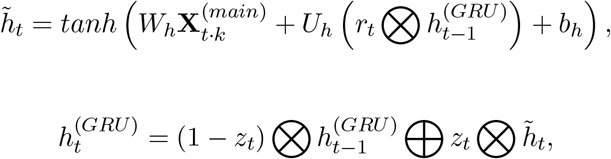

where matrices *W*_*i*_, *W*_*f*_, *W*_*o*_, *W*_*c*_, *W*_*z*_, *W*_*r*_, *W*_*h*_, *U*_*i*_, *U*_*f*_, *U*_*o*_, *U*_*c*_, *U*_*r*_, *U*_*h*_, *U*_*h*_ and vectors *b*_*i*_, *b*_*f*_, *b*_*o*_, *b*_*c*_, *b*_*z*_, *b*_*r*_, *b*_*h*_ are model parameters. *σ* is a sigmoid function,⊗ and ⊕ are pointwise multiplication, pointwise addition, respectively.

For a fully connected layer (*g*^(*dense*)^), we apply a dropout step to limit the dimensions of the outputs, referred to as nodes in the deep learning literature, generated from LSTM and GRU layers and prevent overfitting. The outputs are dropped randomly at a rate to be specified by the users, which we discuss in Section 2.3.3. The number of nodes and the dropout rates for LSTM and GRU layers are tuned as the hyperparameters in the network configurations. The activation function of the fully connected layer is set as the ReLU function to generate the non-negative cumulative confirmed cases and total deaths. Our proposed model, referred to as COVINet, conducts the deep learning process by incorporating county features. The corresponding COVINet is shown in Figure 2.

**Figure 2:**
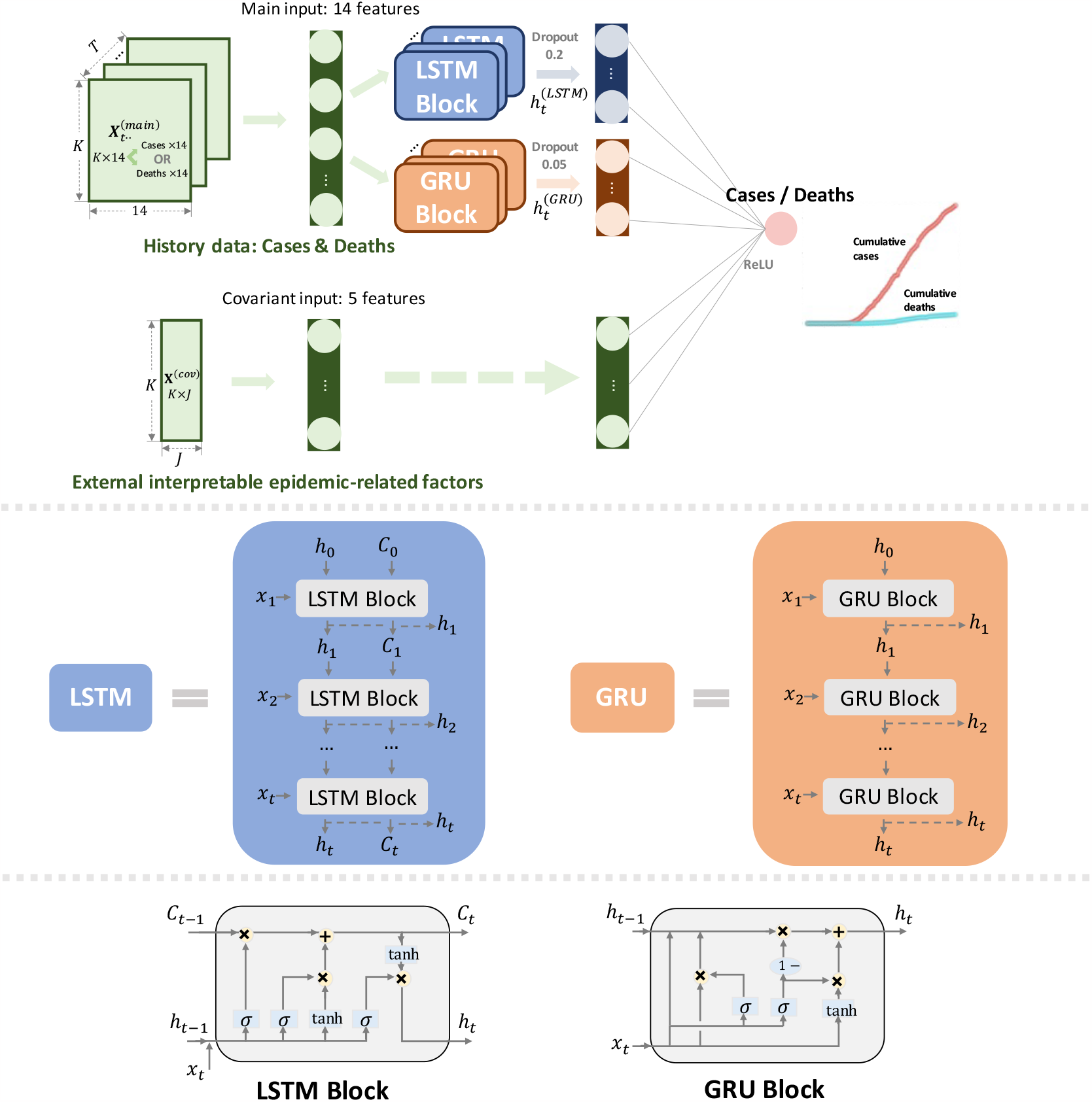
The COVINet combines Long Short-Term Memory (LSTM) and Gated Recurrent Unit (GRU) using *J* (5) county features.

All data involved in the model are min-max normalized within each state before being used. The data from New York City (New York), Macomb (Michigan), Oakland (Michigan), Wayne (Michigan), Cook (Illinois), and Wayne (Illinois), Tarrant (Texas) are normalized separately from the rest of the data in their respective states, because their scales are much larger. This step is found to increase the accuracy of our model and training speed. For unknown data containing the same variables, we use the scales from the training data to transform future epidemic data and then predict the future COVID-19. After obtaining the predicted data, we proportionally restore the predicted cumulative confirmed cases and deaths by reversing the scales.

#### 2.3.2. Training

During the training process, the observed cumulative confirmed cases and deaths in the past fourteen days in each county of the US are used to predict the cumulative confirmed cases and deaths in the 7th day in the future. COVINet is trained to learn the observed patterns of COVID-19 and then to validate the learned patterns, where the accuracy of the models is evaluated by Mean Absolute Errors(MAE_*t*_) and Mean Relative Errors (MRE_*t*_) as validation loss:

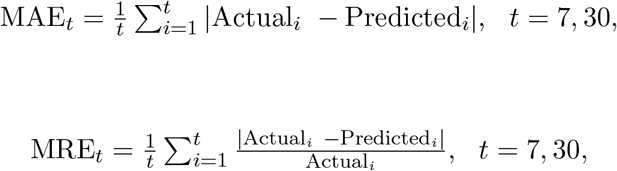

where Actual_*i*_ are the actual cumulative confirmed cases or total deaths at the *i*^*th*^ day and Predicted_*i*_ are the predicted ones at the same corresponding date. The weights of an entire network are estimated by backpropagation through minimizing the loss function (MSE).

We assess the performance of all models through temporal domains. In the comparison between COVINet and the ten CDC models, we utilize data from January 21, 2020, to January 26, 2021, as the training set and data from January 27 to March 23, 2021, as the test set. For additional evaluation, we assess the prediction accuracy for the last eight weeks leading up to March 23, 2023, focusing on the county with the most severe infections in each of the top 10 states.

#### 2.3.3. Tuning the hyperparameters

While building models by LSTM and GRU, we need to tune two hyperparameters to achieve high accuracy. The first one is the number of nodes in LSTM and GRU. We consider 50, 100, and 150 as commonly done [5]. The second one is the dropout rates. We set the range from 0 to 50% with an increment of 5%. The choices of these tuning hyperparameters with the lowest MRE are selected. Specifically, 50 nodes are used for each network in both LSTM and GRU, and the dropout rates are set at 20% and 5% for LSTM and GRU, respectively.

We use the Adam optimizer for model training, and following Kingma and Ba [21], we set *α*=0.001 (step size or learning rate), *β*_1_=0.9, *β*_2_=0.999 (exponential decay rates for the moment estimates), and *ε*=10^*−*7^ for the Adam optimizer. The batch size, i.e., the number of training samples for each iteration, is set as 32. The COVINet model is trained up to 200 epochs. For the learning rate, if the MRE does not decrease for ten consecutive epochs, we reduce the learning rate to its 30% until the MRE decreases or the minimum learning rate reaches 0.00001. The training process is stopped if the MRE does not improve over 40 consecutive epochs.

## 3. Results

### 3.1. Comparison between COVINet and COVID-19 forecast hub models

Table 2 shows the results for the top 10 states and all states in the US for a 7-day prediction. COVINet exhibits outstanding performance with the lowest MAE values among the top 10 states (159.00) and for all states (49.48). Also, COVINet achieves favorable MRE outcomes, with 0.0049 for the top 10 states and 0.0107 for all states. The latter MRE value is close to the minimum MRE of 0.0077 achieved by COVIDhub CDC-ensemble for all states.

### 3.2. Prediction of future trajectories of COVID-19 in the most severe county in each of the top 10 states

The MRE_7_ and MRE_30_ between the observed and projected counts from the day after training periods to March 23, 2023, are computed to assess the accuracy of the temporal prediction for the most severe county in each of the top 10 states, because those hot-hit areas were of the most severe public health interest. Table 3 presents individual MRE_7_ and MRE_30_for those ten counties using COVINet. Overall, the MRE_7_ and MRE_30_ are relatively small, assuring the accuracy of our COVINet model in predicting future trajectories of COVID-19 for the numbers of confirmed cases and deaths for the most severe county in each of the top 10 states.

**Table 3:** MRE_7_ and MRE_30_ of cumulative confirmed cases and deaths using COVINet model with three selected features for each of the ten most severe counties of COVID-19.

| State, County | $MRE_7$ | | $MRE_{30}$ | |
| --- | --- | --- | --- | --- |
|  | Confirmed cases | Deaths | Confirmed cases | Deaths |
| Florida, Miami-Dade | 0.0168 | 0.0087 | 0.0732 | 0.0226 |
| Louisiana, Jefferson | 0.0167 | 0.0080 | 0.0752 | 0.0161 |
| Connecticut, Fairfield | 0.0162 | 0.0103 | 0.0831 | 0.0649 |
| California, Los Angeles | 0.0162 | 0.0089 | 0.0806 | 0.0193 |
| Michigan, Wayne | 0.0169 | 0.0086 | 0.0723 | 0.0241 |
| Pennsylvania, Philadelphia | 0.0176 | 0.0081 | 0.0741 | 0.0199 |
| Illinois, Cook | 0.0166 | 0.0081 | 0.0701 | 0.0263 |
| Massachusetts, Middlesex | 0.0158 | 0.0079 | 0.0745 | 0.0111 |
| New Jersey, Bergen | 0.0177 | 0.0056 | 0.0772 | 0.0177 |
| New York, New York City | 0.0163 | 0.0075 | 0.0769 | 0.0226 |

The 30-day projected trajectories of the cumulative confirmed cases and deaths using the COVINet from August 10, 2022, to March 23, 2023, are presented in Figure From Figure 3, the predicted cumulative confirmed cases from August 10, 2022, to March 23, 2023, are remarkably close to the actual ones for the six counties. The situation is similar in predicting the death counts. The projected values of the confirmed cases for the six counties would increase at a slow rate in the near future.

**Figure 3.:**
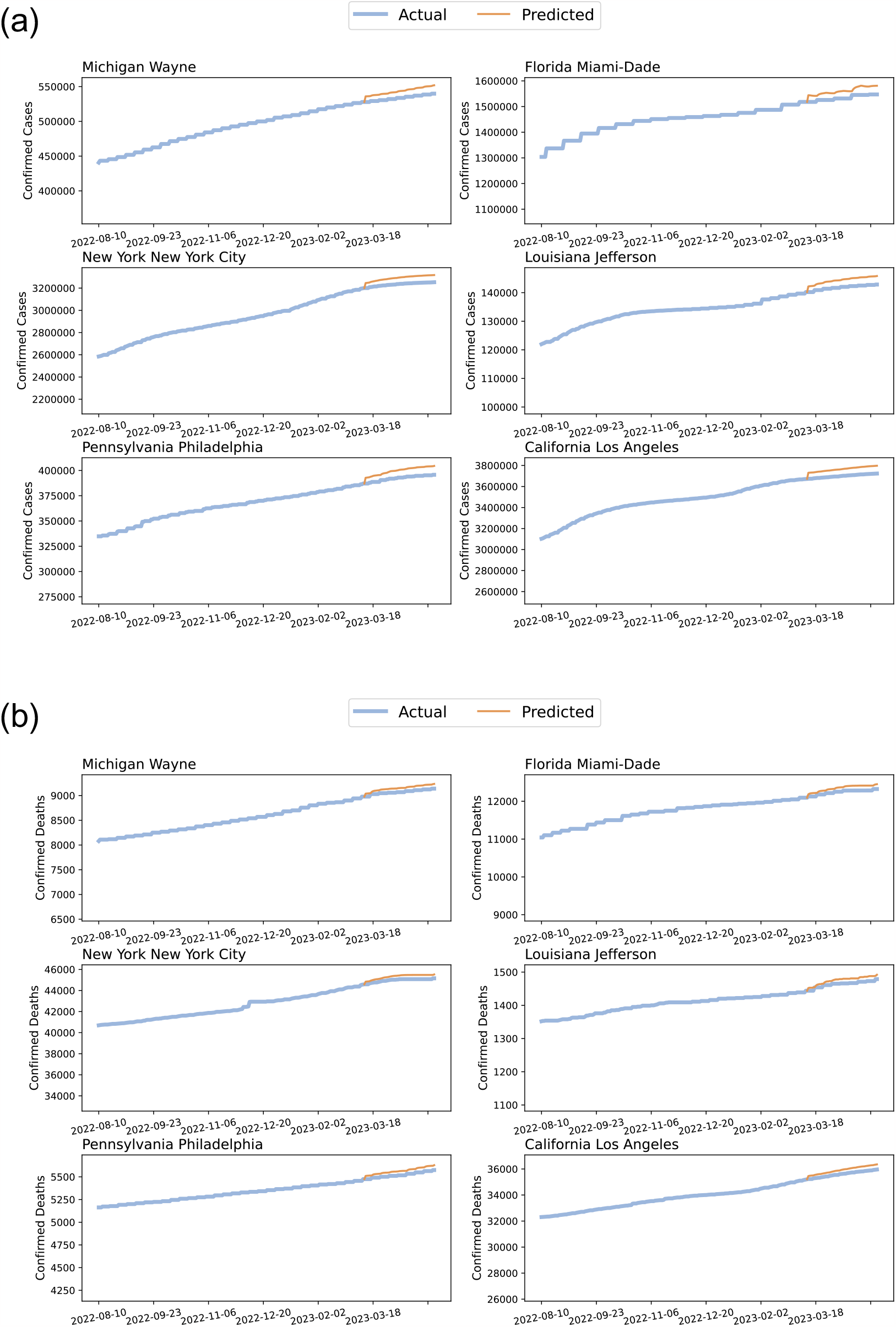
The trajectories of COVID-19 cumulative confirmed cases (a) and total deaths (b) for six counties from August 10, 2022, to March 23, 2023, are displayed. The blue curves indicate the actual cumulative confirmed cases and total deaths, while the orange curves indicate the predicted ones from February 17, 2023, to March 23, 2023.

### 3.3. Feature effects on COVID-19

Our COVINet model incorporates three selected adverse health factors, the longitudes and latitudes of the counties. The weights of longitudes and latitudes are learned from the training county data, where their values are 1.897*×*10^*−*3^ and 4.107*×*10^*−*4^ for the confirmed cases and 2.012*×*10^*−*3^ and 1.021*×*10^*−*3^ for the total deaths, respectively. Accordingly, the Northern and Eastern regions have relatively more confirmed cases, and thus, there are more deaths in the same regions. The maps of the cumulative confirmed cases and total deaths of COVID-19 on March 23, 2023, are presented in Figure 4 and are consistent with our prediction. There are more infected counties in the Northern and Eastern regions.

**Figure 4:**
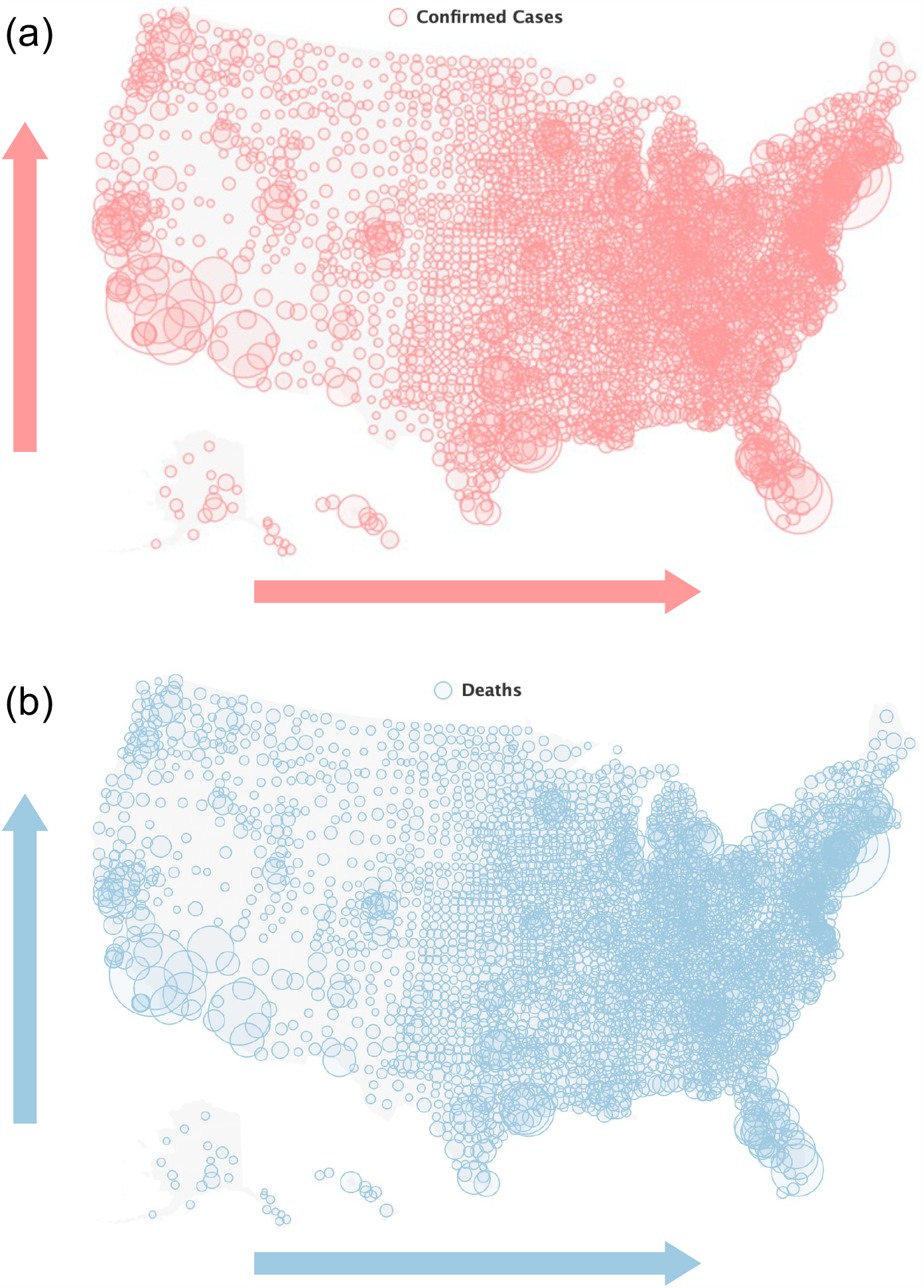
The map of all infected counties. The circle sizes indicate the number of cumulative confirmed cases (a) and deaths (b) on March 23, 2023. The arrows indicate the trend of change in confirmed cases and deaths over longitudes and latitudes.

The weights of the three selected adverse health risk factors are positive for both confirmed cases and deaths. For example, the largest values of weights for confirmed cases and deaths are the traffic volume at 1.783*×*10^*−*3^ and 1.626*×*10^*−*3^, respectively. Specifically, an increase in the traffic volume, severe housing problems, and air pollution would increase both the cumulative confirmed cases and deaths.

To offer insight into the prediction dynamics of COVINet, we vary the levels of the three actional features and present the resulting trajectories of COVID-19 for Los Angeles County, California, as shown in Figure 5. Moreover, for better visibility, we draw the projected trajectories of COVID-19 from March 3, 2023, to March 18, 2023. The impact of the three actional features on COVID-19 in both the cumulative confirmed cases and deaths is visible, depending on the weights of the features. Overall, the number of cumulative confirmed cases and deaths are projected to rise slowly in the following days in Los Angeles County, California. The changes in traffic volume and severe housing problems have a greater impact on the number of confirmed cases and deaths than the changes in air pollution (PM_2.5_), as varying their levels leads to diverging trajectories of confirmed cases and total deaths. The impact of air pollution on the COVID-19 pandemic is relatively slight, as shown by the minimal changes in the cumulative confirmed cases and total deaths across different levels of exposure.

**Figure 5:**
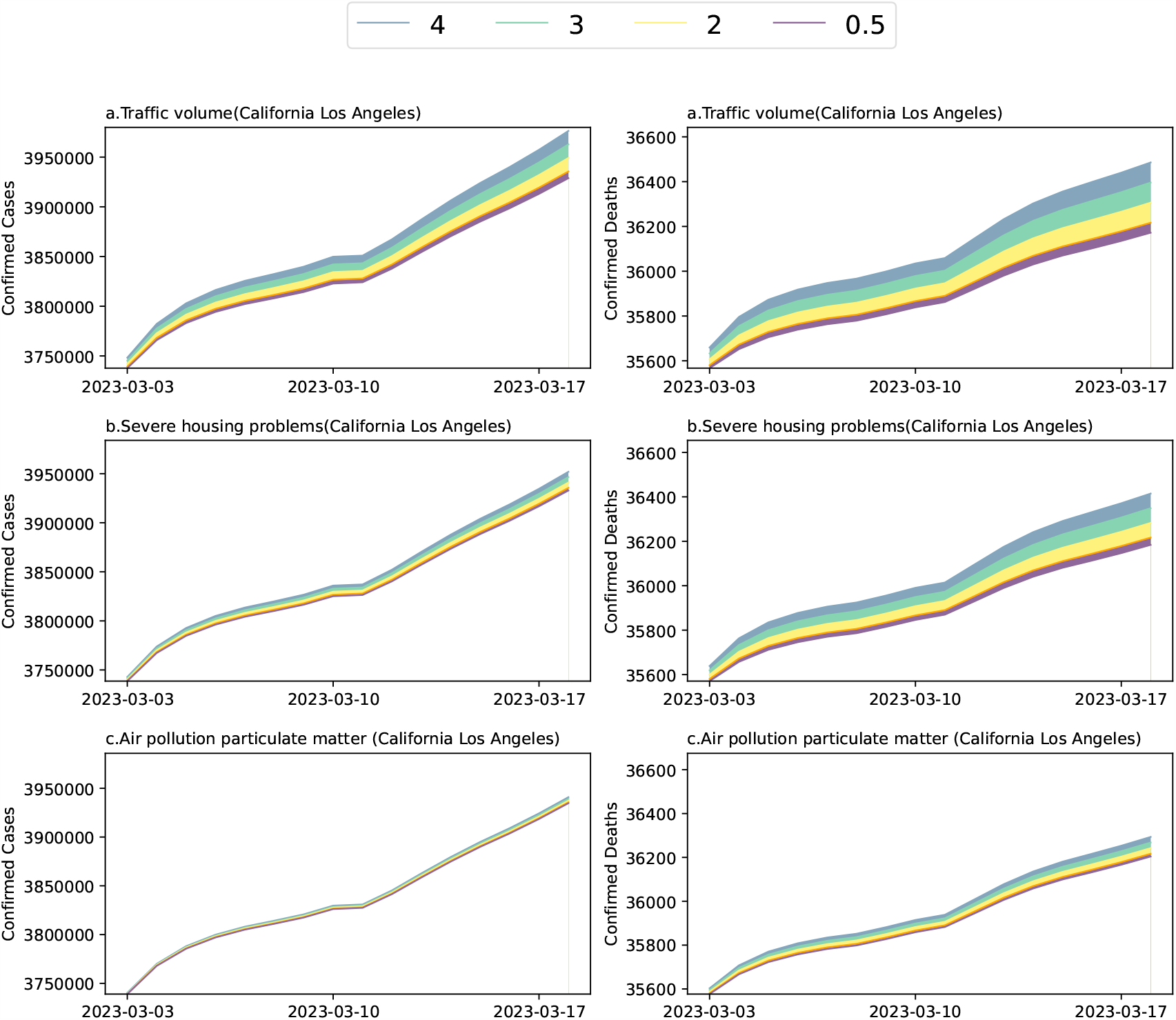
The projected relative trajectories of COVID-19 for Los Angeles County, California, of cumulative confirmed cases and deaths from March 3, 2023 to March 18, 2023. The levels of the three risk factors are changed from 0.5 times to 4 times since January 27, 2023.

## 4. Discussion

Our COVINet is built by deep learning and is shown to be an effective model, which elegantly predicts the cumulative confirmed cases and deaths in US counties. The risk factors that are used in the COVINet provide visible evidence on actionable steps that influenced the trajectories of COVID-19. Thus, COVINet takes advantage of deep learning and the interpretability of risk factors.

LSTM combined with GRU was shown to capture more temporal information, consistent with the work proposed by Dutta et al. [5]. The potential structure of the data that can be captured by using GRU or LSTM alone might be relatively simple. We believe each method alone might not effectively capture the information for accurate prediction. By using both network structures, we can have a more prosperous prediction [5].

To train COVINet, we use the cumulative data (confirmed or death cases) of each county from the previous fourteen days to predict the cumulative data on the 21st day. This time window is chosen because the data from the previous fourteen days contains enough information to capture the trend and the periodicity of the COVID-19 spread. Moreover, the rolling of the data may remove the weekly effect, leading to the model’s better fit of the pattern of COVID-19 trajectories. By rolling the data every day, we can eliminate the weekly effect that may introduce noise or bias to the prediction. For example, the number of confirmed cases might be lower on weekends due to less testing or reporting [7].

In our study, we find that the higher the traffic volume, the higher the risk of COVID-19 spread. A study [44] found that traffic volume was positively associated with COVID-19 incidence and mortality after controlling for population density, income, and others. Traffic volume may reflect the level of human mobility, social contact, and exposure to the virus, which are all crucial for the transmission and outcome of the disease. Moreover, the quality of housing, which may affect the immune system, the respiratory system, and the mental health of the residents, has also been linked to higher COVID-19 infection and death rates. Studies in the US [3] and UK [35] have shown that poor housing conditions, such as overcrowding, dampness, and lack of ventilation, make people more susceptible and vulnerable to COVID-19.

As for air pollution, studies indicate that pre-existing cardiovascular disease could increase the severity of COVID-19 [16, 45], so does the air pollution [12, 13]. The residential proximity to high vehicle traffic at a distance would increase exposure to air pollution and risk of cardiovascular disease (CVD) [6, 9, 25]. However, studies [2, 10, 28] have shown that air pollution has a slight impact on COVID-19 infections, which is in line with the small weights assigned by COVINet to this covariate compared to others.

Overall, if the values of those adverse health factors increase, the trajectories of COVID-19 will be increased accordingly. This might be consistent with the fact that those adverse health factors result in poor health and thus have a high likelihood of increasing the trajectories of COVID-19. Therefore, adverse health factors are expected to differ significantly in the COVID-19 trajectories. As a result of the COVID-19 pandemic, it is a public health matter and an issue of social responsibility.

The estimated weights of covariates in Table 4 align with the variable importance rank obtained from the random forest estimation in Table 1, as well as with the simulated results in Figure 5. The rank for traffic volume, severe housing problems, and air pollution (PM_2.5_) is consistently from large to small. The high degree of consistency in variable importance across different models provides evidence to a certain extent that our model is credible and reliable. There might be other factors that we could consider in building the COVINet. However, we chose to use the three actionable adverse health factors based on a criterion in the random forest, and they may be controllable by local authorities relatively quickly.

**Table 4:** The weights of five adverse health risk factors.

| Factors | Weights (Confirmed Cases) | Weights (Deaths) |
| --- | --- | --- |
| Traffic volume | $1.783 \times 10^{-3}$ | $1.626 \times 10^{-3}$ |
| Severe housing problems | $7.418 \times 10^{-4}$ | $1.236 \times 10^{-3}$ |
| Air pollution | $1.603 \times 10^{-4}$ | $3.184 \times 10^{-4}$ |
| Longitude | $1.897 \times 10^{-3}$ | $2.012 \times 10^{-3}$ |
| Latitude | $4.107 \times 10^{-4}$ | $1.021 \times 10^{-3}$ |

We also take into account the geographical information of infected regions; there could be a link between geographical signals and COVID-19. Our results indicate that higher latitudes have more cases, consistent with previous studies [29, 31]. As the most severe county in the US, the Los Angeles County of California is located in the southwest of the US with the highest number of cases of COVID-19 since 2020. However, for the overall hot-spot areas of COVID-19, approaching north (higher values in the latitude) and east (higher values in the longitude) areas of the US, the more severe counties with higher numbers of cases have been. Accordingly, the same situations apply to the deaths of COVID-19. The majority of severely infected counties are located in the northeast areas of the US.

Our models produce accurate county-level short-term (7-day) and long-term (30-day) predictions of cumulative confirmed cases and total deaths together. More significantly, they are based on measurements routinely surveilled and collected by the local and national authorities, providing actionable information to reduce the spread of COVID-19. COVINet, to some extent, demystifies the black box of deep learning, providing decision-makers with intuitive insights into the impact of health factors on the epidemic. Consequently, it is easy to understand and act by the decision-makers.

## 5. Conclusions

In summary, we built an interpretable and highly accurate prediction model using deep learning for COVID-19. This developed deep learning model can precisely predict the different periods of cumulative confirmed cases and deaths in infected regions. By incorporating the time-invariant factors in deep learning, the accuracy could improve remarkably to predict the trajectories of COVID-19. By analyzing the spread of COVID-19 and adverse health risk factors related to physical and social environments, we can improve the healthcare system for COVID-19.

## Data Availability

We collected the number of cumulative confirmed cases and total deaths fromWe collect the daily numbers of cumulative confirmed cases and deaths from January 21, 2020, to March 23, 2023, for infected counties in the US from the New York Times. The daily cumulative confirmed cases and deaths are collected from health departments and the US CDC, where patients are identified as "confirmed" based on positive laboratory tests and clinical symptoms and exposure. All risk factors are compiled from 2020 annual data on the County Health Rankings and Roadmaps program's official website. In addition, the longitude and latitude of each infected county are collected from Census TIGER 2000. January 21 to May 19, 2020, for counties in the United States from the New York Times, based on reports from state and local health agencies. The county health rankings reports from the year 2020 were compiled from the County Health Rankings and Roadmaps program official website.

https://github.com/nytimes/covid-19-data

https://www.countyhealthrankings.org/reports

## Disclosure statement

### Ethics approval and consent to participate

Not applicable.

### Consent for publication

Not applicable.

### Availability of data and materials

Data are publicly available from the New York Times. The code implementation is available at https://github.com/tingT0929/COVINet-COVID-19.

